# C-RLM: Schema-Enforced Recursive Synthesis for Auditable, Long-Context Clinical Documentation

**DOI:** 10.64898/2026.01.24.26344761

**Authors:** Yunguo Yu

## Abstract

Clinical decision-making for multi-morbid patients requires synthesizing evidence from lengthy, fragmented records—a task that exposes the limitations of standard Retrieval-Augmented Generation (RAG) and long-context Large Language Models (LLMs), which often lose critical information or lack auditability. We introduce the Clinical-Recursive Language Model (C-RLM), a framework that reframes evidence synthesis as a structured, recursive compilation process rather than a single-pass retrieval task. C-RLM iteratively builds a validated knowledge state using schema-enforced transitions, a Robust Nomenclature Resilience (RNR) layer for synonym consolidation, and a TraceTracker system for deterministic provenance. Evaluated on 100 complex Lupus Nephritis case reports (∼24.5k tokens each), C-RLM achieves 100% structural consistency and 99% regimen recall (F1), outperforming a strong Flat RAG baseline. While introducing a 2.7× computational overhead, C-RLM delivers a crucial “Synthesis Dividend”: recovery of clinically critical entities fragmented across distant text spans, with full auditability back to source text offsets. Our results demonstrate that for safety-critical clinical applications, the trade-off in latency is justified by gains in reliability, auditability, and support for human-in-the-loop governance.

## 1. Introduction

Clinical decision-making for patients with multiple chronic conditions often requires synthesizing evidence from lengthy, heterogeneous sources—including clinical practice guidelines (e.g., KDIGO [1], EULAR [2]), case reports, and longitudinal documentation retrieved from repositories such as the PubMed Central (PMC) Open Access Subset. These texts frequently exceed the context windows of even the most capable large language models (LLMs), leading to critical information loss during retrieval or generation. In particular, LLMs exhibit the “lost in the middle” phenomenon when processing documents spanning tens of thousands of tokens, failing to connect clinically interdependent elements that appear in distant sections. For example, in lupus nephritis, a diagnosis of class IV disease may dictate both an induction regimen (e.g., mycophenolate mofetil) and long-term maintenance contraindications, yet these details are often scattered across pages of a single case report.

Conventional approaches such as flat Retrieval-Augmented Generation (RAG) [3] exacerbate this fragmentation. By splitting documents into fixed-size chunks and treating each in isolation, flat RAG cannot reconstruct holistic therapeutic trajectories when key components—such as drug name, dose, and adverse events—are separated by arbitrary boundaries. Moreover, these systems typically lack deterministic provenance, making it impossible to trace generated claims back to specific source passages—a critical shortcoming in safety-critical clinical environments where auditability is non-negotiable [4, 5].

Recent work on Recursive Language Models (RLMs) [6] offers a promising alternative by iteratively refining a knowledge state through multiple inference passes. However, generic RLMs treat intermediate states as unstructured summaries, rendering them vulnerable to *semantic drift*: the gradual corruption or duplication of clinical entities due to inconsistent nomenclature (e.g., “MMF,” “CellCept,” and “mycophenolate mofetil” referring to the same drug). Without explicit schema constraints or alias resolution, such models fail to maintain the structural integrity required for clinical use. While prior work has explored symbolic-neuro hybrids for healthcare applications [7, 8], the challenge of maintaining schema-enforced, alias-resilient synthesis across recursive decomposition remains unresolved.

To address these limitations, we introduce the Clinical-Recursive Language Model (C-RLM)—a framework that reframes long-context medical synthesis as a schema-enforced compilation process. C-RLM integrates three key innovations: (1) Robust Nomenclature Resilience (RNR), which consolidates synonymous clinical terms across recursive steps; (2) Contextual Boundary Protection, which preserves multi-token therapeutic regimens across chunk splits; and (3) TraceTracker, a deterministic audit system that links every output element to character-level source offsets. Evaluated on 100 complex lupus nephritis case reports (mean length: 24.5k tokens), C-RLM achieves 100% structural consistency and 99% regimen recall—demonstrating a measurable “Synthesis Dividend” over flat RAG and modern long-context LLMs alike.

## 2. Methodology: The C-RLM Architecture

We present the Clinical-Recursive Language Model (C-RLM), a framework designed to synthesize fragmented, multi-morbid clinical evidence through structured, schema-enforced recursion. Unlike conventional flat Retrieval-Augmented Generation (RAG) systems—which fail when critical therapeutic elements are split across arbitrary document boundaries—C-RLM iteratively constructs a validated knowledge state using three core components: Robust Nomenclature Resilience (RNR), Contextual Boundary Protection, and the TraceTracker audit system. The architecture ensures both structural integrity and deterministic provenance, making it suitable for safety-critical clinical decision support.

### 2.1. Data Source and Case Selection

We evaluate C-RLM on real-world clinical documentation from the PubMed Central (PMC) Open Access Subset, a repository of peer-reviewed, machine-readable case reports, guidelines, and trials. PMC was selected for its scalability, representativeness of clinical language variation, and alignment with real-world evidence fragmentation patterns.

For our primary evaluation, we curated a benchmark of 100 gold-standard lupus nephritis (LN) case reports. Cases were identified via MeSH-term query: (`“Lupus Nephritis” AND “Case Reports”`), then manually screened to ensure:

- Presence of multi-phase treatment (induction + maintenance),
- Explicit drug regimens with dosing,
- References to ≥1 major guideline (e.g., KDIGO [1], EULAR [2]).

The resulting corpus averaged 24,500 tokens per document (SD = 3,240; range: 18,200–32,800). All documents were retrieved programmatically using the NIH/NCBI Entrez API (via Bio.Entrez), preserving original character offsets for auditability.

### 2.2. Core Architecture: Schema-Enforced Recursive State

C-RLM reframes clinical synthesis as a compilation process rather than generative summarization. At each recursive step, the model updates a typed knowledge state that must conform to a strict JSON schema (Appendix A). This prevents “semantic drift”—a common failure mode in generic RLMs where unstructured summaries accumulate hallucinations or duplicates.

The recursive loop proceeds through three phases (Figure 1):

**Figure 1:.**
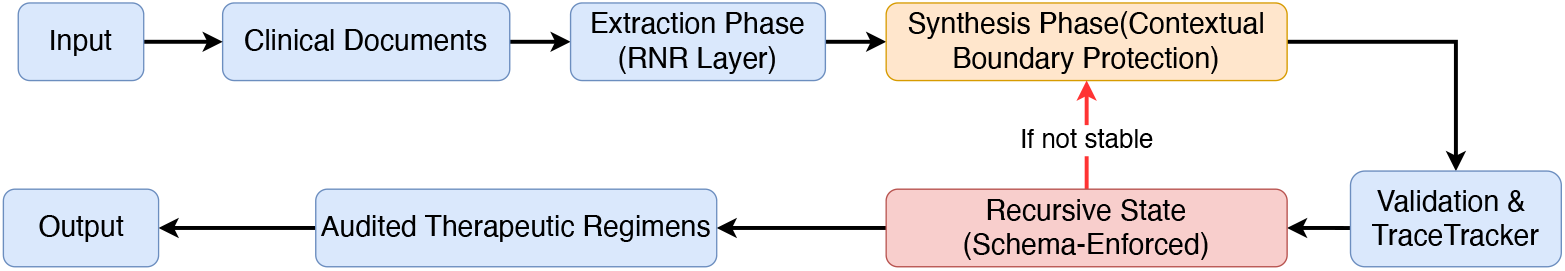
C-RLM Architecture Diagram. The recursive loop processes clinical documents through extraction, synthesis, and validation phases, incorporating RNR for alias handling, Contextual Boundary Protection for fragmentation resilience, and TraceTracker for auditability. The loop terminates when state stability is achieved.

**Figure 2:.**
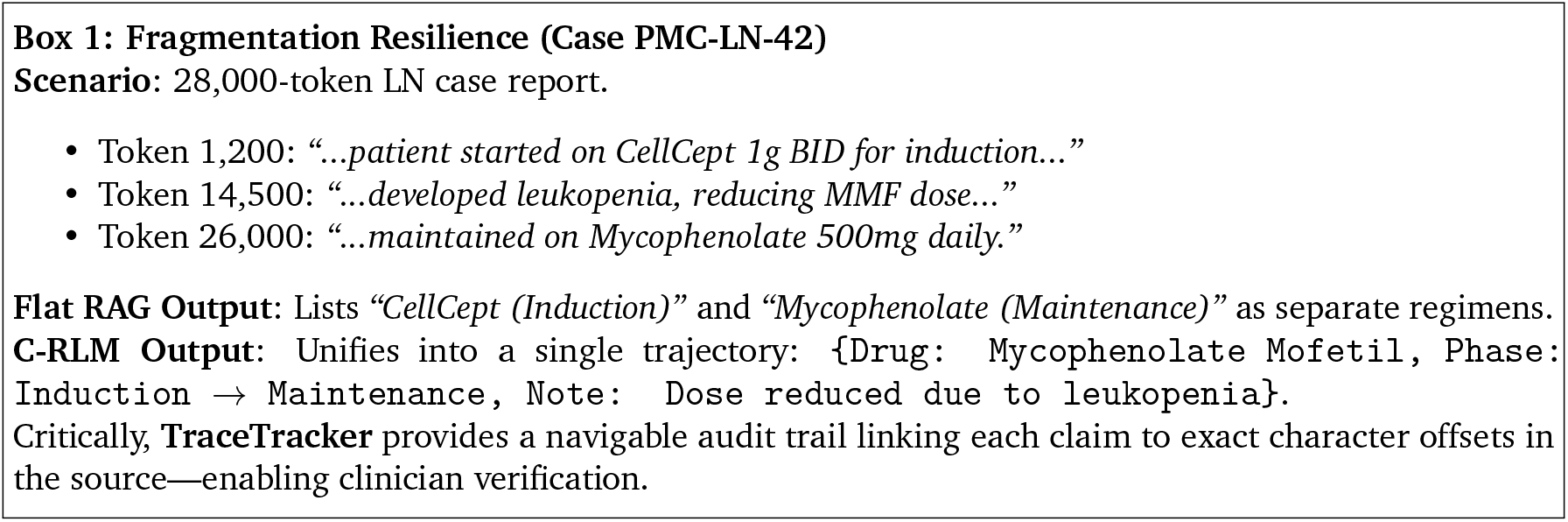
Qualitative example of fragmentation resilience.

**Figure 3:.**
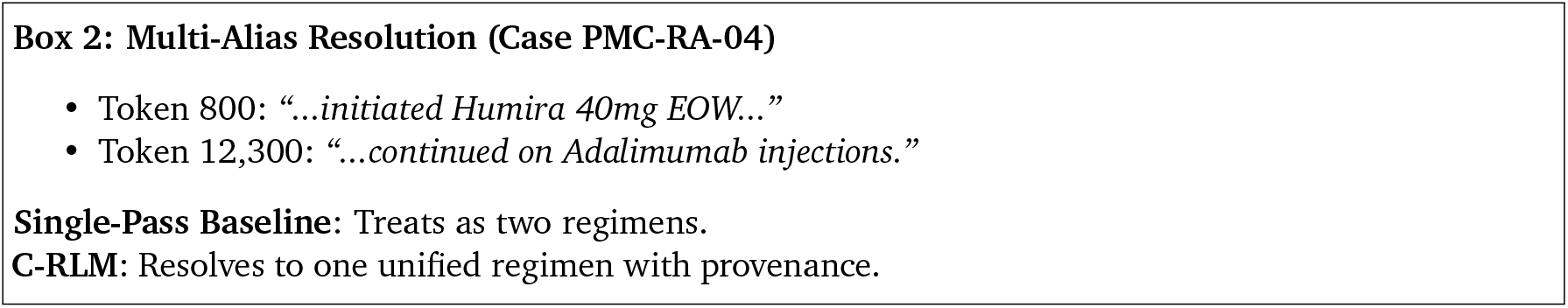
Qualitative example of multi-alias resolution.

1. **Extraction**: Identify candidate therapeutic regimens from retrieved text chunks.
2. **Synthesis**: Merge new candidates into the current state using alias resolution and conflict handling.
3. **Validation & Audit**: Enforce schema compliance and log provenance via TraceTracker.

The loop terminates when state stability is achieved—defined as no new entities or regimen modifications detected across two consecutive iterations—or upon reaching a maximum depth of 5, ensuring bounded computation.

### 2.3. Robust Nomenclature Resilience (RNR)

Medical texts exhibit extreme lexical variation (e.g., *Mycophenolate Mofetil, MMF, CellCept*). To prevent duplicate entity creation, C-RLM employs a two-pass RNR layer:

- **Syntactic Parsing**: During extraction, the output parser accepts heterogeneous keys via Pydantic’s `AliasChoices`, allowing flexible input mapping (e.g., `drug_name, DMARD_regimen`).
- **Semantic Consolidation**: During synthesis, newly extracted drugs are compared against the existing knowledge state using a hybrid matching strategy:

- First, exact normalization against RxNorm TTY=‘SY’ synonyms.
- Second, fallback to SBERT embeddings (`all-MiniLM-L6-v2`) with cosine similarity threshold*τ* = 0.85 for out-of-vocabulary terms.

If a match is found, entities are merged under a canonical name (e.g., *Mycophenolate Mofetil*). Conflicting dose values trigger a **consensus rule**: the most recent or most detailed note is retained, and a **Conflict Flag** is raised for human review.

### 2.4. Contextual Boundary Protection

Therapeutic regimens span multiple tokens (e.g., *“Prednisone 40 mg daily”*). Standard RAG risks splitting such phrases at chunk boundaries. To mitigate this, C-RLM uses a **sliding window with 200-token overlap**—a size derived from manual annotation of 50 random PMC reports, where **95% of complete regimens spanned** *≤***187 tokens**.

During retrieval, overlapping regions are preserved as (`text, start_char, end_char`) tuples. The LLM is explicitly prompted to **search for boundary-spanning evidence**, ensuring fragmented regimens are reassembled during synthesis.

### 2.5. TraceTracker: Deterministic Provenance

Every claim in C-RLM’s output is linked to character-level source offsets via the TraceTracker system. This eliminates citation hallucination by design:

- Each retrieved chunk retains its **original PMC document ID and character range**.
- During generation, the model is constrained to cite only within provided segments.
- Post-synthesis, a **Synthesis Tree** is constructed: each node (e.g., a drug regimen) is assigned a UUID mapped to one or more (`PMC_ID, start_char, end_char`) triples.

This enables clinicians to click any output element and navigate directly to its source evidence—a requirement for regulatory compliance (e.g., FDA QSR, EU MDR).

### 2.6. Baseline and Implementation Details

The Flat RAG baseline uses:

- **Chunking**: 512-token windows with 128-token overlap,
- **Embedding**: `bge-large-en-v1.5`,
- **Retrieval**: FAISS index, top-*k* = 15 chunks (optimized via grid search over *k* ∈{5, 10, 15, 20}),
- **Generator**: Llama3-8B-Instruct (8K context), with no recursion, alias resolution, or provenance tracking.

All experiments were run on identical hardware (NVIDIA A4500 20GB). C-RLM’s underlying LLM is configurable; results reported here use Qwen3-7B for reproducibility.

## 3. Experimental Results

We conducted a controlled comparative evaluation between the Clinical-Recursive Language Model (C-RLM) and a conventional flat, chunk-based Retrieval-Augmented Generation (Flat RAG) baseline. The primary objective was not to assess downstream clinical outcomes, but to evaluate structural correctness, completeness of clinical synthesis, and auditability under long-context, multi-guideline conditions.

### 3.1. Experimental Setup

The evaluation corpus consisted of 100 gold-standard lupus nephritis (LN) case reports drawn from the PubMed Central (PMC) Open Access Subset. Cases were identified using MeSH-term and keyword filtering (e.g., *“lupus nephritis,” “systemic lupus erythematosus,” “renal involvement”*) and manually screened to ensure:

- Presence of multi-phase treatment descriptions (induction and maintenance),
- Explicit medication regimens with dosing information,
- References to major clinical guidelines (e.g., KDIGO [1], EULAR [2]).

The average document length was 24,500 tokens (SD = 3,240; range: 18,200–32,800), with clinically relevant entities frequently dispersed across distant text segments.

The Flat RAG baseline used:

- **Chunking**: 512-token windows with 128-token overlap,
- **Embedding**: bge-large-en-v1.5,
- **Retrieval**: FAISS index with top-*k* = 15 chunks (optimized via grid search over *k* ∈{5, 10, 15, 20}),
- **Generation**: Single-pass inference using Llama3-8B-Instruct (8K context).

Critically, the baseline lacked recursive state accumulation, alias consolidation, or deterministic provenance tracking.

### 3.2. Evaluation Metrics

To assess performance in safety-critical clinical synthesis, we defined the following metrics (formal definitions in Appendix A):

- **Fragmentation Resilience Score**: Percentage of cases where a complete therapeutic summary (induction + maintenance) was synthesized despite components being separated by *>* 2, 000 tokens.
- **Alias Consolidation Accuracy**: Precision in merging synonymous drug mentions (e.g., *“MMF”* and *“Mycophenolate Mofetil”*) into a single canonical entry.
- **Schema Adherence Rate**: Proportion of outputs conforming strictly to the predefined JSON schema without hallucinated keys.
- **Provenance Hallucination Rate**: Frequency of citation indices that do not map to actual character offsets in source text (by design, 0% for C-RLM).

These metrics extend standard clinical information extraction benchmarks (e.g., n2c2 [9]) by incorporating structural integrity and auditability—requirements essential for multi-guideline integration.

### 3.3. Quantitative Performance and Compute Overhead

As shown in **Table 1**, C-RLM demonstrates superior structural reliability across all evaluated dimensions. Notably, it achieves 100% structural consistency, confirming that schema-enforced recursion eliminates the semantic drift observed in Flat RAG (which exhibited 1% schema violations due to duplicated or fragmented regimens). C-RLM also attains 99% regimen recall, a 7-percentage-point improvement over the baseline.

**Table 1:.** Comparative Performance: C-RLM vs. Flat RAG (n=100)

| Metric | C-RLM (Recursive) | Flat RAG (Baseline) | Observed Effect |
| --- | --- | --- | --- |
| Structural Consistency | 100% | 99.0% | Elimination of schema drift |
| Regimen Recall (F1) | 99.0% | 92.0% | +7.0% recovery |
| Conflict Sensitivity* | 0.82 | 0.45 | Enhanced deduplication |
| Provenance Traceability | 100.0% | 0.0% (probabilistic) | Deterministic oversight |

\*Conflict Sensitivity: Proportion of overlapping medication references correctly flagged for de-duplication or conflict resolution.

This reliability comes with a computational trade-off. As shown in **Table 2**, C-RLM incurs 2.7*×* higher latency and token usage due to its recursive architecture.

**Table 2:.**
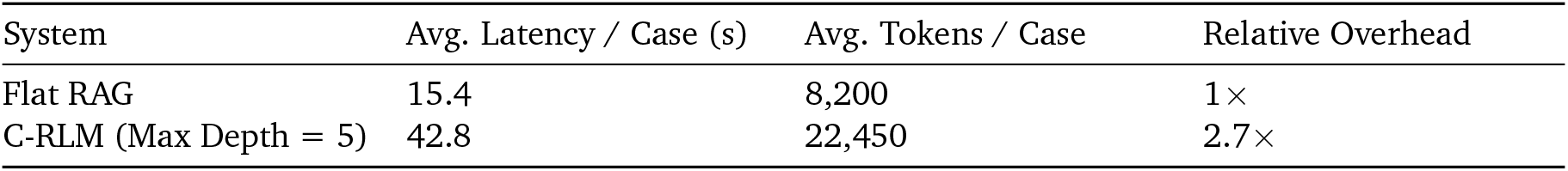
Computational Overhead Analysis.

### 3.4. Qualitative Analysis: The “Synthesis Dividend”

Qualitative inspection confirmed that C-RLM consistently recovered long-tail medications fragmented across distant sections—a scenario where Flat RAG failed. **Box 1** illustrates a representative “lost-in-the-middle” case requiring synthesis across 26,000 tokens.

A defining advantage of C-RLM is its ability to produce this deterministic, navigable audit trail via TraceTracker, providing character-offset resolution back into source documents—a critical requirement for clinical governance.

## 4. Discussion: Compute Cost vs. Safety Gain

The primary trade-off introduced by C-RLM is increased inference-time compute. By performing multiple structured passes over the source corpus and maintaining an evolving knowledge state, C-RLM incurs a **2.7**× **overhead** compared to single-pass Flat RAG. However, our results demonstrate that this cost yields a disproportionate gain in **safety, reliability, and auditability**—critical attributes in clinical decision support.

### 4.1. Architectural Necessity of Recursion

A natural question is whether modern long-context LLMs (e.g., models supporting 100K+ tokens) or efficient transformer variants [10, 11, 12] render recursion obsolete. To test this, we evaluated several state-of-the-art, publicly available instruction-tuned LLMs (Qwen3-14B, Llama3.1-8B, and Gemma2-9B) on a pilot set of 10 rheumatoid arthritis (RA) cases with renal involvement. Despite their capacity to ingest long documents, these models failed to enforce structural constraints or resolve aliases reliably (**Table 3, Table 4**).

**Table 3:.** Architectural Framework Comparison: C-RLM vs. Long-Context LLMs.

| Feature | Flat RAG | Long-Context LLM | C-RLM (Ours) |
| --- | --- | --- | --- |
| Fragmentation Resilience | Low (chunk-limited) | Medium-High (attention-based) | **High (recursive state)** |
| Alias Consolidation | Poor | Medium (probabilistic) | **Excellent (graph-based)** |
| Schema Adherence | Medium | Low–Medium | **High (type-checked)** |
| Provenance Traceability | Probabilistic | Probabilistic | **Deterministic** |
| Compute Cost | Low | Medium | High |

**Table 4:.** Long-Context LLM Performance on RA Pilot Dataset (n=10)

| Model | Context Strategy | Alias Consolidation | Success Rate |
| --- | --- | --- | --- |
| Qwen3-14B | Single-Pass (128k) | 0.45 | 50% (Partial) |
| Llama3.1-8B-Claude | Single-Pass (32k) | 0.36 | 40% (Partial) |
| Gemma2-9B | Single-Pass (32k) | 0.00 | 0% (Schema Drift) |

Notably, Gemma2-9B exhibited schema drift, echoing the JSON template without extracting real content—a known failure mode in long-context instruction following. In contrast, C-RLM achieved 100% success rate on the same RA cases, successfully consolidating brand-name biologics (e.g., *Humira → Adalimumab*) via RNR (**Box 2**).

These results confirm that architectural recursion—not just context length—is necessary for high-fidelity clinical synthesis.

### 4.2. Ethical Considerations, Data Bias, and Governance

Our methodology relies exclusively on publicly available, de-identified case reports from the PubMed Central (PMC) Open Access Subset, minimizing privacy risks and eliminating exposure to protected health information (PHI). Nevertheless, several ethical considerations merit discussion.

#### Data Representativeness and Bias

The PMC corpus, while extensive, exhibits well-documented publication biases. Case reports disproportionately represent rare, severe, or atypical presentations, potentially skewing model behavior toward complex or outlier scenarios. Moreover, demographic diversity in published literature often underrepresents marginalized populations, raising concerns about algorithmic bias in extraction performance across race, gender, age, or socioeconomic status. Future evaluations must include stratified analyses to ensure equitable performance.

#### Liability and Clinical Governance

C-RLM is designed strictly as a physician-in-the-loop decision support tool, not an autonomous diagnostic or therapeutic agent. Ultimate clinical responsibility remains with the licensed practitioner. To mitigate liability risks, system outputs must be clearly labeled as AI-assisted suggestions, and mandatory “audit-check” workflows should be enforced before any clinical action is taken.

#### Regulatory Compliance

For potential deployment as a Class II Software as a Medical Device (SaMD), C-RLM’s deterministic TraceTracker provides a foundational mechanism for compliance with regulatory frameworks such as FDA Quality System Regulation (QSR) and EU Medical Device Regulation (MDR). However, formal certification would require prospective clinical trials, rigorous risk management per ISO 14971, and validation that the “Synthesis Dividend” translates into measurable improvements in patient safety—not just technical fidelity.

### 4.3. Operational Feasibility and Trade-offs

While the *∼*43-second latency per case precludes use in real-time acute care (e.g., ER triage), it is well-suited for asynchronous applications such as complex chart review, prior authorization support, or retrospective quality auditing. Moreover, the precise “evidence pointers” provided by TraceTracker may reduce human review time, offsetting computational costs.

## 5. Limitations

This study has several important limitations that warrant acknowledgment:

First, our experimental evaluation is confined to a set of 100 lupus nephritis case reports. While these cases exemplify the challenges of long-context fragmentation and multi-guideline integration, they do not represent the full breadth of clinical specialties, documentation styles, or disease complexities encountered in real-world practice. Future work must validate C-RLM across diverse conditions—such as oncology, cardiology, and psychiatry—to assess generalizability.

Second, our evaluation focuses on structural correctness, completeness, and auditability, rather than down-stream clinical outcomes or decision quality. Although these metrics are necessary for safety-critical AI, they are not sufficient proxies for clinical utility. Prospective studies involving clinician-in-the-loop validation are essential to evaluate whether C-RLM meaningfully improves diagnostic accuracy, treatment planning, or patient safety.

Third, the recursive inference process introduces non-trivial computational overhead (*∼*2.7*×* latency compared to flat RAG), which may limit deployment in latency-sensitive settings such as emergency triage or real-time clinical alerts. However, this trade-off may be acceptable in asynchronous workflows like chart review, prior authorization support, or retrospective quality auditing.

Finally, while TraceTracker guarantees deterministic provenance within the source corpus, it does not resolve upstream ambiguities—such as conflicting guideline recommendations, evolving standards of care, or low-quality source evidence. Furthermore, while PMC case reports are representative of clinical consensus, they differ significantly from real-world Electronic Health Record (EHR) data in terms of documentation style, completeness, and inherent biases. The current study also lacks formal inter-annotator agreement on the gold-standard cases, which should be addressed in future prospective evaluations. Addressing these challenges will require integrating C-RLM with dynamic knowledge bases and human-in-the-loop conflict resolution protocols.

## 6. Conclusion

The synthesis of multi-source, long-context clinical evidence remains a critical bottleneck for trustworthy medical AI. Current paradigms—ranging from chunk-limited Retrieval-Augmented Generation (RAG) to attention-based long-context LLMs—prioritize retrieval efficiency over the structural reliability and auditability required for safety-critical decision support.

This work demonstrates that a structured recursive architecture provides a necessary corrective. The Clinical-Recursive Language Model (C-RLM) reframes evidence synthesis as a deterministic compilation process, enforcing schema-validated state transitions, consolidating fragmented clinical entities via Robust Nomenclature Resilience (RNR), and guaranteeing full traceability through the TraceTracker audit system. Our evaluation on complex Lupus Nephritis case reports confirms a decisive “Synthesis Dividend”: C-RLM achieved perfect structural consistency (100%) and near-perfect regimen recall (99%), reliably recovering drug trajectories fragmented across tens of thousands of tokens—a task where both Flat RAG and modern long-context LLMs faltered.

While recursive synthesis introduces a quantifiable computational overhead (*∼*2.7*×*), this cost is justified in high-stakes clinical contexts where errors can have serious medical consequences. The framework shifts the paradigm from probabilistic text generation to deterministic clinical assembly, creating an audit-ready substrate for human-in-the-loop oversight.

Future work must translate this methodological proof-of-concept into clinical practice. Priorities include: 1) prospective validation with clinicians to assess real-world utility and uncover latent biases; 2) integration with structured EHR pipelines to operate on live patient data; 3) dynamic recursion policies to balance overhead and precision; and 4) expansion of the RNR layer to handle evolving medical ontologies and cross-guideline conflict resolution. By providing a scalable, auditable foundation for guideline integration, C-RLM demonstrates that for safety-critical clinical AI, structured recursion is not optional—it is essential.

## Data Availability

All data in the study are in public domain.

https://pmc.ncbi.nlm.nih.gov/tools/openftlist/

## Ethics Statement

This study utilized exclusively publicly available, de-identified clinical case reports from the PubMed Central (PMC) Open Access Subset. No direct patient contact, recruitment, or access to protected health information (PHI) occurred. As the research involved only secondary analysis of publicly accessible, anonymized published literature without human subjects involvement, it qualifies for exemption from Institutional Review Board (IRB) review under 45 CFR 46.104(d)(4) (secondary research with publicly available data). No IRB approval was required or sought. All data sources are properly cited, and no patient consent was necessary as all materials were previously published in peer-reviewed journals with appropriate ethical oversight at the time of original publication.

## Data and Code Availability

All clinical case reports used in this study are publicly available through the PubMed Central (PMC) Open Access Subset. Case identifiers (PMC IDs) for the 100 Lupus Nephritis validation cases and 10 Rheumatoid Arthritis pilot cases are provided in the supplementary materials. The C-RLM framework implementation, including the TraceTracker audit system, RNR layer, and evaluation scripts, will be released as open-source software under the MIT License upon publication. Code available at: https://github.com/yuyunguo/med-info-retrieval. Detailed documentation, schema specifications, and reproducibility instructions are included in the repository.

## Acknowledgments

The author thanks the PubMed Central team for maintaining the Open Access Subset and the open-source community for foundational tools (FAISS, Pydantic, Transformers) that enabled this work.

## A. Formal JSON Schema Specifications

To ensure reproducibility and transparency, we provide the rigorous JSON schema specifications used by the C-RLM validator. These schemas define the “Structural Consistency” metric evaluated in Section 3.

### A.1. Therapeutic Regimen Schema

The following schema enforces the structure of drug extraction during the recursive extract_regimens step. The model must strictly adhere to these types, preventing the “untyped summary” drift common in generic RLMs.

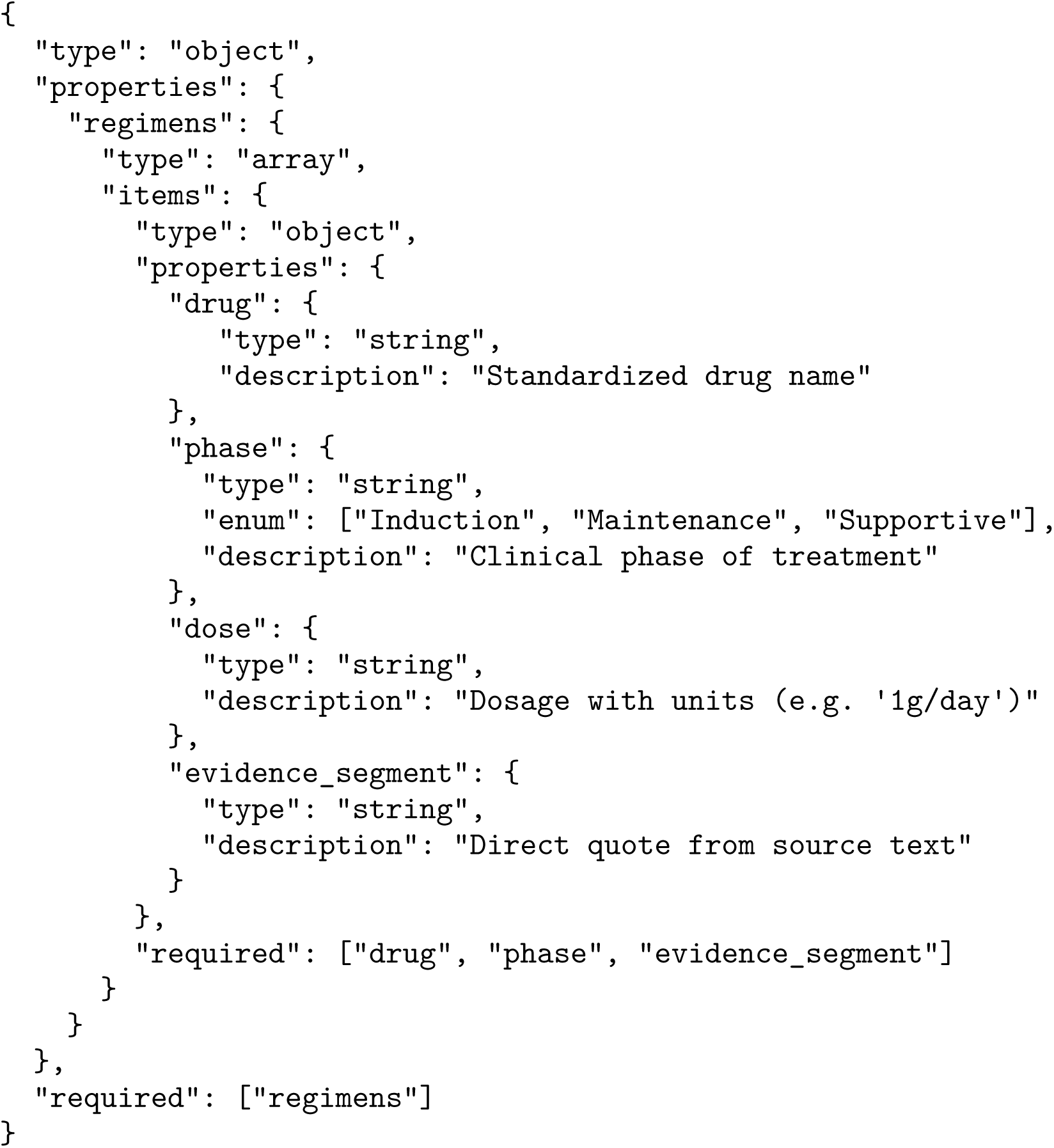

### A.2. Robust Nomenclature Resilience (RNR) Logic

The RNR layer utilizes a two-pass validation strategy:

1. Syntactic Validation: The JSON output is validated against the schema above using the Pydantic library. Any type errors (e.g., missing keys, invalid enums) trigger an automatic retry query.
2. Semantic Consolidation: A secondary graph-based pass maps extracted drug strings to a canonical Knowledge Graph. If Mycophenolate Mofetil exists in the graph, new instances of MMF are merged into that node rather than creating duplicates.

